# The impact of the COVID-19 pandemic on mental health and well-being of people living with a long-term physical health condition: a qualitative study

**DOI:** 10.1101/2020.12.03.20243246

**Authors:** A Fisher, A Roberts, A.R. McKinlay, D Fancourt, A Burton

## Abstract

**Background:** The COVID-19 pandemic and associated restrictions caused major global disruption. Individuals with long-term physical health conditions (LTCs) are at higher risk of severe illness and often subject to the strictest pandemic guidance, so may be disproportionally affected. The aim of this study was to qualitatively explore how living with a LTC during the COVID-19 pandemic affected people’s mental health and wellbeing.

**Sample and methods:** 32 participants, mean age 57 (SD 13) years, 66% female and 72% white British, who reported having LTCS (most commonly cancer, respiratory conditions or cardiovascular diseases), participated in telephone/video call interviews based on a semi-structured topic guide. Key themes and subthemes were determined using deductive and inductive thematic analysis.

**Results:** There were four overarching themes specific to living with a LTC. These were 1) high levels of fear and anxiety related to perceived consequences of catching COVID-19, 2) impact of shielding/isolation on mental health and wellbeing, 3) experience of healthcare during the pandemic and 4) anxiety created by uncertainty about the future. Fourteen subthemes were identified, including concerns about accessing essential supplies and the importance of social support. Individuals who lived alone and were advised to shield could be profoundly negatively affected.

**Conclusions:** This study found that there were a number of aspects of living with a LTC during the pandemic that had a significant impact on mental health and well-being. There should be focus on how best to provide practical and social support to people with LTCs during a pandemic, particularly if they have to shield or isolate.

## Introduction

The COVID-19 pandemic caused global disruption, with nearly every country introducing social distancing measures. Individuals with long-term physical health conditions (LTCs) like diabetes, hypertension, and respiratory conditions are particularly susceptible to severe illness if infected by the virus that causes COVID-19 (Sars-Cov-2) (Apicella et al., 2020; Li et al., 2020; Sanyaolu et al., 2020) and therefore were often subject to the strictest restrictions. Indeed, if considered extremely clinically vulnerable, people living with LTCs in the UK were advised to ‘shield’ at the beginning of the pandemic (not leaving their home/garden and avoiding social contact) (PHE, 2020). Understanding the impact of the challenges faced by people living with LTCs during the pandemic is crucial in determining who has been negatively affected, what support they may need during and beyond the pandemic, and how they could be supported should future pandemics occur.

Quantitative surveys suggest that the impact of lockdown and shielding had negative effects on mental health and well-being for some, but not all, people with LTCs. An Office for National Statistics (ONS) report estimated that that 35% (of 2.2 million people) living in the UK experienced poorer mental health as a result of shielding (ONS, 2020). The COVID-19 Social Study, a longitudinal survey of 70,000 respondents, found that those with LTCs were more adherent to UK Government lockdown guidelines than those without, but reported higher levels of stress, depression and anxiety throughout (Fancourt et al., 2020). Other longitudinal surveys also suggest that the pandemic restrictions had a negative impact on mental health and well-being in individuals with LTCs, although some showed no difference compared to the general population (Alonzi et al., 2020; Bargon et al., 2020; Pierce et al., 2020). However, long-term physical and mental health problems often co-occur (e.g. (Yohannes et al., 2010)), so the extent to which these findings were a result of the pandemic and associated restrictions is unclear. Indeed, in-depth research has not been undertaken to understand how individuals with LTCs experience a pandemic and what unique challenges they may face.

Qualitative research can elucidate such experiences and help untangle seemingly discrepant findings (Teti et al., 2020). Qualitative methodologies have also proved invaluable in understanding the mental health impacts during other infectious disease emergencies, like the spread of Ebola (Johnson & Vindrola-Padros, 2017). However, at the time of writing, there was limited qualitative data from individuals with LTCs collected during the COVID-19 pandemic. In an early-phase survey of 7039 individuals with long-term respiratory conditions, qualitative analysis of free text suggested that perceived vulnerability to infection, limited access to healthcare and necessities, uncertainty about the future and inadequacy of the UK Government response were having extremely negative impacts on mental health (Philip et al., 2020). Free text responses are valuable, but limited by the inability to probe. A qualitative interview study of 30 young adults with Type 1 diabetes conducted in India in June 2020 found that participants had low awareness of their elevated risk of severe illness from COVID-19 and low awareness of COVID-19 symptoms or preventative measures (Pal et al., 2020). Further, in a qualitative study of the impact of COVID-related delays in 31 adults awaiting kidney transplantation in Australia, participants described disappointment/devastation, concerns about vulnerability, additional burdens (like financial) and high levels of stress created by uncertainty (Guha et al., 2020). However, these studies were specific in their scope. Therefore, the aim of this study was to qualitatively explore how living with a LTC during the COVID-19 pandemic in the UK affected people’s mental health and wellbeing.

## Methods

This study was part of a wider investigation of the psychological and social impacts of the COVID-19 pandemic: the COVID-19 Social Study www.covidsocialstudy.org. Participants in the qualitative arm of the COVID-19 Social Study, a large-scale exploration of the impact of the pandemic on a number of subgroups, were recruited by circulating advertisements widely to third sector organisations, via the MARCH network (www.marchnetwork.org) and research team networks via social media and email cascade. For inclusion in this study, participants had to be adults (≥18 years), be fluent in spoken English and have been diagnosed with any condition they perceived to be a long-term physical health condition. Participants for this study were purposely sampled to capture different age groups, genders, ethnic backgrounds and types of LTCs using targeted advertisements via social media, the COVID-19 Social Study website and newsletter (3,919 subscribers). Ethical approval was provided by UCL Ethics Committee (Project ID 14895/005) and all participants provided informed written consent prior to interview. Methods and results are presented in line with the Consolidated Criteria for Reporting Qualitative Research (COREQ) checklist (Tong et al., 2007).

### Qualitative interviews

Participants completed a pre-interview demographic questionnaire reporting age, gender, ethnicity, education, marital status and details of LTCs. LTCs were coded into 12 categories for descriptive purposes. Interviews were conducted via telephone or video call by four female Health Psychology/Social Science postdoctoral researchers experienced in qualitative interviewing (AR, AM, RC, SE). The interviews followed a topic guide (presented as Supplementary Material) which was developed using existing theories on behaviour change (Michie et al., 2011), social networks and health (Berkman et al., 2000) and stress, health and coping (Eriksson, 2017). Questions were designed to explore how the COVID-19 pandemic has affected mental health and wellbeing, social lives, and social behaviours.

### Data Analysis

Interviews were audio recorded and transcribed by a professional transcription service with a UCL data sharing agreement. Transcripts were anonymised and checked for accuracy before being transferred to Nvivo 12 for analysis. Thematic analysis was conducted following the steps outlined by Braun and Clarke (Braun & Clarke, 2008). AR undertook line-by-line coding of 23 transcripts during ongoing data collection using an initial deductive coding framework based on the interview schedule. AM and AR double-coded three transcripts and compared codes to ensure consistency of approach. AR continued coding transcripts using a modified coding framework based on content described by participants as coding progressed (inductive approach). Results were presented to a group of qualitative COVID-19 Social Study researchers, who provided feedback and comments on the analysis over several weeks. AM applied the coding framework to the final 9 transcripts then AF, AM and AB reviewed, defined and named the themes, selected the quotes and produced the report. Themes and sub-themes are presented with illustrative quotes, with participant gender, age range (in years) and brief description of LTC to preserve anonymity.

## Results

32 participants took part and participant characteristics are presented in **Table 1**. Individuals were aged 32-75 years and the majority were female (n=21/66%) and white British (n=23/72%), but a wide range of demographic groups were represented. LTCs are reported in full in **Table 2**, but most commonly reported were cancer (n=12/38%), respiratory conditions (n=10/31%) and cardiovascular diseases (CVD) (n=9/28%). Nine participants (28%) also reported a co-occurring mental health condition. Interviews were between 31 and 150 (mean 65) minutes long.

**Table 1.** Participant characteristics.

| **Demographics** |  |
| --- | --- |
| **Age (years)** |  |
| Range | 32-75 |
| Mean (SD) | 57 (13) |
| **Gender n (%)** |  |
| Female | 21 (66) |
| Male | 11 (34) |
| **Ethnicity, n (%)** |  |
| White British | 23 (72) |
| White - Other | 3 (9) |
| Indian | 2 (6) |
| Mixed ethnic groups | 2 (6) |
| Black - Other | 2 (6) |
| **Marital status, n (%)** |  |
| Married/Civil Partnership | 16 (50) |
| Divorced/separated | 7 (22) |
| Single (never married) | 8 (25) |
| Widowed | 1 (3) |
| **Education, n (%)** |  |
| GSCE (or equivalent) | 3 (9) |
| A-levels (or equivalent) | 5 (16) |
| Undergraduate degree | 12 (38) |
| Postgraduate degree | 12 (38) |
| **Employment, n (%)** |  |
| Employed | 9 (28) |
| Unemployed | 19 (60) |
| Furloughed | 3 (9) |
| Studying | 1 (3) |
| **Living situation, n (%)** |  |
| Live alone | 9 (28) |
| Live with others | 23 (72) |

**Table 2.** Long-term health conditions (LTCs)

| **Health conditions, n (%)<sup>a</sup>** |  |
| --- | --- |
| Respiratory (e.g. asthma, COPD) | 10 (31) |
| Cardiovascular (e.g. high blood pressure, heart disease) | 9 (28) |
| Musculoskeletal (e.g. osteoporosis, osteoarthritis) | 8 (25) |
| Cancer (in remission) | 7 (22) |
| Current cancer diagnosis (incl. advanced/incurable cancer) | 5 (16) |
| Diabetes | 5 (16) |
| Autoimmune (e.g. rheumatoid arthritis) | 5 (16) |
| Neurological (e.g. Parkinson's Disease, Multiple Sclerosis) | 4 (13) |
| Renal (e.g. kidney condition, interstitial cystitis) | 2 (6) |
| Inflammatory Bowel Disease (e.g. Crohn's) | 2 (6) |
| Organ removal/transplant (e.g. spleen removed, liver transplant) | 2 (6) |
| Comorbid mental health condition (e.g. anxiety, depression, schizophrenia) | 6 (19) |
| <sup>a</sup> percentages do not equal 100% due to comorbidity among participants |  |

Themes and subthemes are summarised in **Table 3**. The most commonly emerging themes relating specifically to living with a LTC were 1) high levels of fear and anxiety related to perceived consequences of catching COVID-19, 2) major impact of shielding/isolation on mental health and wellbeing, 3) experience of healthcare during the pandemic and 4) anxiety created by uncertainty about the future. Other more general themes about the impact of the pandemic that were common across COVID-19 Social Study groups will be the subject of future papers.

**Table 3.** Impact of COVID-19 pandemic on people with LTCs Summary of themes *(and subthemes)*

|  |
| --- |
| **1) High levels of fear and anxiety related to perceived consequences of catching COVID-19** |
| *Heightened awareness of risk due to LTC* |
| *Fear of hospitalisation, ventilation and death* |
| **2) Impact of shielding/isolation on mental health and wellbeing** |
| *Concerns about access to essential supplies* |
| *Immediate social network strongly influenced experience of shielding* |
| *Reliance on others and loss of independence difficult* |
| *Feeling safe versus feeling isolated* |
| **3) Experience of healthcare during the pandemic** |
| *Move to remote interaction with healthcare services mostly positive* |
| *Mixed views of attending healthcare settings in person* |
| *Postponement of non-essential treatments* |
| *Threat of disruption to cancer treatments created stress and anxiety* |
| **4) Anxiety created by uncertainty about the future** |
| *Impact of pandemic on treatment access with progression of LTC* |
| *Fear of restrictions being lifted and plans to continue to isolation* |
| *Not having an end in sight* |
| *Acceptance most protective for mental health* |

### 1) High levels of fear and anxiety related to perceived consequences of COVID-19 infection

#### 1.1 Heightened awareness of risk due to LTC

Participants described fear and anxiety caused by awareness of the potential implications of catching COVID-19. Some described a heightened sense of risk, coupled with an understanding of the need to protect themselves:

> *“There’s been some degree of added stress, I suppose, because it became relatively clear, relatively early, that [COVID-19’s] something that really, given my circumstances, I should be careful not to catch.” (male, 30-39, cancer)*

However, heightened awareness of risk did not always lead to additional worry, with some participants discussing elements of chance or fate in the outcome:

> *“My medical condition, I’ve had it all my life so if that places me in a higher risk category, you’ve just got to do your absolute best to not catch it and then after that it’s sort of in the lap of the Gods really isn’t it? So it didn’t really upset me or stress me or worry me” (female, 40-49, respiratory condition)*

#### 1.2 Fear of hospitalisation, ventilation and death

Other participants described the specific risks that COVID-19 infection could lead to, including long-term health problems, risk of hospital admission, ventilation, and dying, and acknowledged the negative impact of this on mental health:

> *“I don’t want to catch it, [I have] already experienced being on a ventilator before for 17 days and I never want that experience again*… *(female, 60-69, cancer, CVD)*
>
> *“I’m terrified of getting this virus, because I know that if I get it, it probably is the end of me. My lungs are not good…I don’t want to die in hospital and I don’t want to have that intubation and sedation” (female, 70-79, respiratory condition)*
>
> *“I’m really scared of getting [COVID-19]…I think I probably would not survive if I got it, so I’m trying to keep away from it…when I heard about [COVID-19], I just automatically went, oh my God, I’m going to die. It wasn’t great” (female, 30-39, CVD, respiratory condition)*

### 2) Impact of shielding/isolation on mental health and wellbeing

The COVID-19 restrictions imposed on the participants’ lives led to a number of negative consequences for mental health. Many were in the extremely clinically vulnerable group and therefore advised to shield, or had made the decision to isolate. This led to specific concerns, as well as discussion of factors that alleviated these worries.

#### 2.1 Concerns about access to essential supplies

Participants discussed the ability to access essential supplies (e.g. food, medical supplies) as having strong influences on mental health. Those who found it difficult to access food and medical supplies were fearful and anxious:

> *“they ended up cancelling my vulnerable status on their [online shopping] system, cancelling my delivery slot And now the next delivery slot is three weeks away. So, what am I meant to do? How the hell am I going to cope now? I haven’t got three weeks’ worth of food (male, 30-39, cancer, respiratory condition)*
>
> *“…the pharmacy’s not providing any service of delivery to vulnerable people, and the GP’s are arguing with me, to say, why are you ordering medication a week before it’s due date? Or whatever, but I can’t order one item at a time, who’s going to go and pick it up, for a start? And when it runs out, it’s going to run out, and I’m not going to have anything to replace it” (female, 50-59, multiple conditions)*

Participants reported issues around receiving the necessary shielding certification that would allow access to (e.g.) priority status online supermarkets:

> *“I didn’t get a letter [from the Government/National Health Service (NHS)] for ages because they weren’t able to identify me and the therapy from that essential health data. So, yes it was a bloody nightmare to be honest” (female, 40-49, cancer)*

Those who had Government certification/other shielding arrangements (like social and neighbourhood networks) in place acknowledged the positive impact of this on their mental health:

> *“I registered very early with the supermarket vulnerable list, I think, that the government was sharing with them. And I think because I got my shielded letter, very early on I got an email from [supermarket] saying, you’re on the vulnerable list and we’ll prioritise slots. So we’ve been very lucky that we actually managed to get slots when we need” (female, 30-39, autoimmune condition)*

Others spoke about how they stockpiled supplies to reduce anxiety:

> *“I actually made sure that I had an extra stock [of inhalers]. I put an extra repeat prescription in before the lockdown and I’ve just put another one in*..*” (female, 40-49, cancer)*
>
> *“I suppose I panicked a bit and I got a delivery thinking I’d better be prepared. I do have a certain amount of food in anyway because living alone, I’ve got to be prepared for a bad cold or something…I thought oh, a couple of months, I’d better order all the tinned stuff…” (female, 70-79, neurological and musculoskeletal conditions)*

### 2.2 Immediate social network strongly influenced experience of shielding

In addition to the impact of social networks in supporting shielding, participants described polarised experiences of whether they felt supported by friends and family members to shield. Those who felt supported described the positive impact:

> *“Most of the time [the family are] in the house…Everybody’s on the same page…Maybe if I hadn’t had gone through everything I went through last year and when kids see you poorly and what have you, and it’s cancer, so it’s the C word, isn’t it? It’s made everybody think on the same wavelength…it’s had its moments…But as a family, we’ve done well with it to be honest” (female, 50-59, cancer)*

However, some described challenges that arose from having to justify their need to shield. This led to worries about offending others, arguments, and had an extremely detrimental impact on mental health:

> “*Without sounding harsh, my sister just didn’t get this idea of me needing to isolate. So, she kept trying to come up and see us. We were relying on her to bring us food and things like that. But she just kept hanging around*..*… I think that was probably one of the tougher ones because I think from her point of view, she just seemed to think that I just wanted her to not be around, whereas this is the advice I’d been given” (male, 30-39, cancer, respiratory condition)*
>
> *“[my son] was living at home…he left during the lockdown because I had the letter from the NHS saying I was vulnerable. At the beginning, his girlfriend came and then she went. And he wanted her to come back and I had to say, no…so he left…It was awful…I felt absolutely devastated… The other [son] had gone a few months before he’d gone. I was left on my own, my partner had left me [last year] after 22 years…I blamed myself. I didn’t know. I was very down, very depressed” (female, 50-59, cancer, respiratory condition)*

Some participants also reported feeling aggravated by friends and family who they believed were not following the social distancing rules relevant to them and this putting strain on relationships:

> *“we’ve got friends who are in their 70s, and they’re so gung-ho about the whole thing. We’ve had arguments with them to get them to stay in, because they’re all really social…we had a hell of a job persuading them not to do that because of what’s happened, because it’s all just dissolved into a shambles” (male, 60-69, neurological condition)*

### 2.3 Loss of independence and reliance on others

Although social networks were important, participants discussed the need to rely on others and loss of independence as difficult:

> *“I was especially jealous of my girlfriend, who didn’t have to shield. She made sure that before she starts work in the morning, she goes for a walk for at least 45 minutes to an hour… She also had to do the grocery shopping. And there was once or twice during lockdown that I needed my prescriptions to be refilled. She had to do that. So, in a sense, I felt overly reliant on her…*.. *So yes, that was difficult (male, 40-49, blood condition)*

The difficulty in relying on others resulted in one participant going out to shop despite guidance against it:

> *“I don’t like relying on people. I hated to have to ring people up. At the very beginning, lots of people were getting in touch…Then it tailed off a little bit and I don’t like ringing people and asking for help…I just felt really guilty for it. I just thought, I won’t bother anybody, I’ll go and do it myself” (female, 50-59, respiratory condition)*

### 2.4 Feelings of safety versus isolation

Some participants reported that, despite missing friends and family, the enhanced feeling of safety and security through lockdown was reassuring and protective for mental health and a worthwhile trade-off:

> *“we’re in lockdown, it’s not great, but…I think the sense of relief kind of outweighed any frustrations really……I’m just happy to proceed like this until we see what’s going to happen…” (female, 30-39, bowel condition)*

However, others found not seeing friends and family one of the most challenging aspects to deal with:

> *“I’m desperately worried about [my mother-in-law]. She’s got serious Alzheimer’s. She’s alone, she’s not seen any of her family for months and she’s just sitting in a room. She’s starving to death because she’s not eating any more. She’s going to die at some point probably in the next six weeks at this rate. And it’s possible that we’re not going to be able to see her. It’s horrendous” (male, 40-49, autoimmune condition, CVD)*

Some participants who were shielding and living alone suffered particularly with social isolation. Two disclosed that the extent of the social isolation (in combination with other factors related to the pandemic that were detrimental to mental health) led them to contemplate/attempt suicide:

> *“because you’re stuck at home and if you don’t talk online to somebody or over the phone to somebody then you’re on your own and you’re just going to go downhill. There’s that fear that I could get back to being suicidal…I’ve had periods where I have felt very low and struggling and very alone, and I think living on your own is just shit. People that are part of a couple or a family and whatever have got other people around” (female, 50-59, neurological condition)*

Others discussed loss of physical contact as particularly difficult:

> *“there’s the desire to hug and shake hands, just human contact sometimes…I miss a little bit of human contact obviously” (male, 50-59, CVD)*

### 3) Experience of healthcare during the pandemic

Experiences of the impact on healthcare and treatments were commonly discussed, with both positive and negative aspects reported.

#### 3.1 Move to remote interaction with health services mostly positive

Many participants described how their healthcare consultations had changed to telephone/video appointments during the pandemic, and this was often regarded as a more convenient alternative:

> *“there’s been a big push to change some things, like phone clinics and video clinics…it’s been brilliant. And my care is split over three different hospitals, so rather than spend 45 minutes each way on the Tube to get into my hospital, wait in a busy clinic and then get back…It’s so much easier to just be able to phone*…*” (female, 30-39, bowel condition)*
>
> *“I have a routine discussion with the oncology unit at the hospital, which has been by telephone ever since I moved down here. I’ve had one face-to-face meeting with the oncologist and he said, well you have to have telephone routine checks, and I said that’s fine with me, I don’t have to drive over, park the car, hang around for an hour or two. So this actually works better…” (male, 70-79, cancer)*

Although one participant felt it wasn’t as satisfactory and another talked about missing the reassurance normally gained from a physical assessment and having concerns that it was not as medically rigorous:

> *“my clinic appointments have been over the phone. The bit that I’ve missed out on is somebody physically checking my lymph nodes…” (female, 40-49, cancer)*

#### 3.2 Mixed views of attending healthcare settings in person

Participants described varying emotions regarding attending healthcare services during lockdown. Some felt that the precautions taken by healthcare professionals (wearing personal protective equipment, promoting hand-washing, triaging appointments) alleviated anxiety, and they felt safe and reassured:

> *“I felt okay about [going for a blood test], I know the GP surgery quite well and they’re all very friendly and competent as far as I can see…if there had been about 20 people in the waiting room, I wouldn’t have been too happy about that, but they’re actually managing the flow of people extremely well” (male, 70-79, cancer)*

Others described feeling anxious initially, however for those attending regularly this anxiety diminished:

> *“The first time I went, I don’t know because the nurses have not been tested. And I thought I’m shielding at home and yet I’m coming here and you’re giving me treatment intravenously and you’ve not been tested and that didn’t sit right with me…But now I’m fine, I go every three weeks It’s fine” (female, 50-59, cancer)*

Some felt extremely anxious about catching COVID-19 during hospitalisation for their LTC, particularly if accident and emergency (A&E) admission was required, whilst others felt the risk of this was low:

> *““In my head, if I went to A&E and went into the hot side with Coronavirus, that’s just a death sentence” (male, 30-39, cancer, respiratory condition)*
>
> *“I didn’t feel any anxiety about going into hospital because most hospitals are designated as red zones and green zones. Certainly, all the nurses etc. wore masks, gloves and all the rest of it. But I had excellent care and I wasn’t at any point worried that I would actually catch [COVID-19] in hospital” (female, 60-69, respiratory condition)*

Some participants described evaluating the perceived seriousness of health problems they were experiencing versus the potential risk of catching COVID-19 in deciding whether to access healthcare services:

> *“the night I had the chest pain, it mentally went through well, I’ll give it another 10 minutes and see. If it hasn’t gone, I will go to hospital. So I wasn’t just going to sit there and think I’m going to put up with chest pain just because I might get COVID-19 if I go to hospital. I was sensible enough to realise that really, the more pressing thing’s getting what seemed to be a heart attack sorted than worrying about a theoretical risk of getting [COVID-19]” (female, 60-69, respiratory condition)*

However, others reported real reluctance to engage with healthcare services. One described not attending an appointment at the hospital because of their concerns about the risk of contracting COVID-19, and another delayed help seeking for suspected cancer recurrence:

> *“I had possible signs of a resurgence of the cancer. I had a lump come up on my neck. And we were already in lockdown at that point…And so I put it off, of course, as we usually do, and seeing if it would go away. But two weeks later it was still there…And it was clear I had to do something because I was tearing myself apart with panic*…*” (female, 70-79, cancer in remission)*

#### 3.3 Postponement of non-essential treatments

Some participants had non-essential treatments or appointments postponed. Some were in agreement with this decision, while others experienced disappointment (despite understanding the necessity of it):

> *“I should have seen my neurologist in January…but that got cancelled. I contacted [the hospital] and got an answer phone, then I was told that the neurologist would be getting in touch and she never was…. A bit disappointed and I do feel like I think what I’ve got is a pretty serious condition, but it’s obviously regarded as not that important at the moment…but I can understand why” (male, 60-69, neurological condition)*
>
> *“I was supposed to have a surgical review with a view to having surgery this year, but obviously that’s all stopped. The review was cancelled. Surgery will not be any time soon. It can wait, but it’s also something that’s disappointing. But I do understand that there will be a massive backlog now and there are many more urgent things that need sorting” (female, 40-49, respiratory condition)*

#### 3.4 Fear of disruption to cancer treatments created stress and anxiety

Participants undergoing cancer treatment spoke about the stress and anxiety caused by threat of disruption to treatment. One participant was undergoing chemotherapy with curative intent:

> *“There was a bit of stress at some point because…[the oncologist] was hinting, basically, that potentially they would have to cut my treatment short to a degree…I guess the consequence of that is that you increase the risk of things coming back and all that, so it wasn’t ideal. But 15 days later, when I went back for my next cycle, that was out of the window and things have improved in [the hospital]” (male, 30-39, cancer)*

Another was receiving treatment for incurable cancer that was intended to limit cancer progression and prolong her life, and was relieved that her treatment was not affected:

> *“my treatment’s carried on, they were talking about cancelling it, but they didn’t…I was glad, because you never know. It’s under control at the minute, but I know things can change, so I was glad mine carried on.” (female, 50-59, cancer)*

### 4) Anxiety created by uncertainty about the future

Whilst a number of participants recognised that the experience of living with a LTC may have somewhat prepared them to cope with uncertainty, they still described a number of issues specific to the pandemic that were challenging for mental health:

> *“[uncertainty’s] something that I wouldn’t say I’m an expert in dealing with, but I’m very experienced in dealing with it. But it doesn’t make it any easier. But it seems to be something I’ve had to deal with, at least on the back burner, all my adult life” (male, 50-59, diabetes)*

#### 4.1 Impact of pandemic on treatment access with progression of LTC

For some participants, the pandemic exacerbated their worries about future treatment, progression of their LTC, and ability to access healthcare in the future if their health deteriorated:

> *“there is the underlying anxiety, as well, about how will I be treated in the future?*…*The healthcare worry is obviously a lot more intense, because if I need to go back into treatment, I will have to be isolated to a much greater degree, and will be much more dangerous…it’s always been there, but [now] it’s an increased anxiety” (female, 50-59, cancer)*
>
> *“healthcare is my main priority, that really worries me, that I’m not going to get the same level of treatment as I was getting before, because there won’t be sufficient money around, and a lot of services will be cut” (female, 50-59, multiple conditions)*

This worry seemed less pronounced in those with cancer, who emphasised that their main worries about the future were about the cancer, and the pandemic had not changed that:

> *“[cancer’s] always a worry, really. I think I’m very much of the opinion that it will come back at some point, and that’s kind of what the numbers show. I’m not particularly emotionally worried about that…that’s not changed because of this situation” (female, 40-49, cancer)*

One participant, who was currently receiving cancer treatment, described how they tried not to look beyond completion of this:

> *“if I’ve got any worries about the future, in the ranking, cancer comes a clear first and the pandemic is some way behind that …I’ll need to process what it means to the future of me, in the future, once the treatment is gone” (male, 30-39, cancer)*

#### 4.2 Fear of restrictions being relaxed and plans to continue isolating

Many participants discussed anxiety around relaxation of guidelines and plans to continue shielding or isolating even if/when restrictions were lifted:

> *“Even now shops are opening and stuff. I’m too anxious to go out and about …. I’ll just keep monitoring what’s going on and make my own decision as to when I feel it’s safe for me to get back into the big world” (female, 50-59, neurological condition)*
>
> *“And I personally feel that it will take a while before I would feel comfortable going out. So, regardless of what the government says, I will be preferring my own guidelines” (female, 60-69, cancer in remission, CVD)*

However, participants undergoing active cancer treatment felt less concerned about relaxation of the lockdown, since their treatment meant they would have to isolate anyway:

> *“I’m not concerned so much, because…I’m going to be isolating for longer than everybody else, so when the country starts to reopen, I’m going to be here. And if there’s a second wave, I’m going to witness that from where I sit here. I’m not going to be part of the second wave” (male, 30-39, cancer)*

### 4.3 Not having an end in sight

Despite some reporting they would electively remain in isolation after restrictions were relaxed, several participants mentioned that the uncertainty around how long the pandemic would last as particularly difficult:

> *“how long’s the virus going to be floating around for? How long have we got to take these precautions?*…*I can’t anticipate whether we’re talking weeks, months or a couple of years and the long-term effects are going to be floating around. The anxiety I’m sure will lessen but I think it’s going to be there for a good while” (female, 50-59, neurological condition)*

Some described a sense of loss or grief in not knowing when (or if) social lives would return to normal:

> *“It’s fine for the moment, but, obviously if I think I’m never going to see the Royal Ballet again, I can get quite tearful. And it’s things like dancing, we do dance quite a lot…obviously we can dance together, but it’s not the same as going dancing, so yes. It’s a grieving for how quickly those things come back” (female, 50-59, cancer)*
>
> *“it’s almost like a sense of mourning for the life that we’ve lost. Because it’s hard to see how it can ever go back to being quite what it was before…that grieves me a bit” (female, 70-79, cancer in remission)*

Many participants also expressed that they felt that normal life would not resume until a vaccine had been found and placed their hopes for a return to normality on a successful vaccine:

> *“…. This has crippled the world and will continue to do so until a vaccine is found. That’s quite a daunting prospect on normality” (female, 40-49, respiratory condition)*
>
> *“to me, the only thing that will stop this is a vaccine…that’s the real hope” (male, 50-59, CVD)*
>
> *“I think the uncertainty of it and just not being sure how it’s going to end. And when they also say things like oh, we may never get a vaccine, for f**k sake, and then what? Do I never go out again? I think that’s it. Probably the worst bit” (female, 30-39, CVD, respiratory condition)*.

### 4.5 Acceptance most protective for mental health

Despite the challenges of uncertainty, many participants described that they had found trying to accept the situation as the most protective coping mechanism for their mental health:

> *“I guess it’s just a massive thing that’s outside anyone’s control, so you just have to adapt. You have to be very flexible and adapt to it” (male, 50-59, diabetes)*
>
> *“what’s the point of railing against it? All you’re doing is making yourself upset. The situation is the situation…you’ve just got to accept that it will be what it will be and you make the best of it” (female, 70-79, neurological and musculoskeletal conditions)*

## Discussion

This study found living with a LTC during the COVID-19 pandemic had a major impact on multiple different aspects of mental health and wellbeing. Whilst some experiences of people living with LTCs mirrored those of people without (Brooks et al., 2020), this study identified a range of themes with subthemes that were particular to living with a LTC during a pandemic. Themes included anxiety created by perceived consequences of catching COVID-19, the impact of shielding/isolation, impact on healthcare and anxiety created by uncertainty about the future. Each of these findings has clear implications for the continued management of the COVID-19 pandemic but also preparation for future epidemics, with these implications discussed below.

Participants commonly reported that anxiety about the perceived consequences of catching COVID-19 were having very negative impacts. This is in line with a free text analysis of 7039 participants with respiratory disease, a qualitative analysis of patients awaiting kidney transplant (Guha et al., 2020; Philip et al., 2020) and a large quantitative survey (Fancourt et al., 2020). Individuals with certain LTCs are at elevated risk of worse outcomes if they catch COVID-19, and awareness of this appears high. This highlights the importance of providing helpful information on strategies to reduce risk, along with balanced and accurate messaging about level of risk for people with LTCs during epidemics, to help alleviate anxiety. In a rapid review of the psychological impact of quarantine in previous infectious disease outbreaks, lack of adequate information about level of risk was a key psychological stressor (as was fear of infection), so the authors recommend giving as much reliable information as possible as a mitigating strategy (Brooks et al., 2020).

Building on this, it is also important that support is available to ensure that any guidance introduced to reduce risk (like social distancing/isolating) are feasible and tolerable. For example, participants reported not always being able to access essential supplies whilst shielding, with concerns over this access playing a key role in determining how negatively lockdown guidance (particularly shielding/isolation) was experienced. This echoes findings from previous studies, including the study of participants with respiratory disease and a rapid review of the psychological impact of quarantine (Brooks et al., 2020; Philip et al., 2020). These challenges led to some participants going out against advice to access essential supplies. Quantitative data from the ONS Shielding Behavioural Survey suggest that the need to access essential supplies was the most common reason for people breaking shielding (ONS, 2020). Our participants reported practical factors, like timely receipt of Government shielding status, as having a major impact on whether they were eligible for priority schemes to receive deliveries of supplies, providing some clear targets for support for the continuing COVID-19 pandemic and future similar situations.

Social support to shield or isolate was also a factor in influencing how detrimental the experience of shielding/isolation was to mental health in our sample. In over 1 million people in the UK who felt able to shield, 74% reported that regular contact from friends and family was a main factor in enabling them to adhere (ONS, 2020). For participants in our study, immediate social networks were the most important social connections and this raises awareness of how strongly immediate social support influences the experience of people with LTCs. Acceptance, support and understanding for the needs of the person with the LTC to shield/isolate supported coping, but participants also discussed challenges including their perceived loss of independence and reliance on others. Loss of independence was related to worsening mental health in a large longitudinal cohort prior to the pandemic (Albanese et al., 2020) and considering the negative impact of loss of independence on mental and well-being is part of National Institute for Health and Care Excellence (NICE) guidance for working with older adults (NICE, 2015). Considering how to balance the impact of loss of independence, whilst ensuring people with LTCs get the support they need when shielding/isolating is an important area for future research.

In line with the aforementioned free text study of individuals with respiratory conditions, participants in the current study very commonly discussed impact on healthcare as one of the most prominent factors affecting their mental health and wellbeing during the COVID-19 pandemic. In our study there were encouraging positives; for most, the move to remote consultations was viewed as a convenient and acceptable alternative. Many of the factors that were causing fear and anxiety appeared based on perceptions (i.e. fear of *potential* disruption to cancer treatments, or fear of being admitted to COVID-19 zones in hospital) did not materialise in practice. Indeed those who were initially fearful and then had to attend regular appointments found their fears diminished. However, there were occasions of participants reporting weighing up presentation of potentially seriously symptoms with fear of attending healthcare. This is concerning as it could have implications for the development of diseases and burden on the healthcare service if such diseases are diagnosed later on. Our findings reflect quantitative data showing decline in healthcare attendance during the pandemic. For example, data from the UK NHS suggest that A&E attendance was 29% lower during pandemic restrictions than in the same month the previous year (NHS England, 2020). The US Centre for Disease Control estimated that 41% of people with one underlying health condition, and 55% with two or more, were delaying or avoiding medical care due the COVID-19 pandemic (compared to 30% without) (Czeisler et al., 2020). Future research should explore how people with LTCs can be reassured that it is safer to attend healthcare settings than delay presenting symptoms.

Finally, a consistent theme in qualitative studies conducted in people with LTCs during the pandemic (including ours) was uncertainty about the future having a negative impact on mental health (Guha et al., 2020; Philip et al., 2020). Whilst people without LTCs may share these concerns, the reasons identified in our study were specifically related to having LTCs (e.g. fear of their conditions progressing and treatments not being available due to COVID-related pressure on health services). Participants reported that the most effective way they had found to mitigate the negative impact of the pandemic-related uncertainty on their mental health was to develop acceptance. Acceptance Commitment Therapy incorporates this central idea of acceptance and has shown early promise in people with LTCs (Graham et al., 2016) so could be worth exploring for pandemic-related distress.

### Strengths and limitations

To the best of our knowledge, this was the first study to qualitatively explore the impact of the COVID-19 pandemic and associated restrictions in the UK on the mental health and well-being of people with LTCs. In this paper we focussed on the emergent themes that were specific to living with an LTC. However, there were other more general themes discussed so this study alone may not reflect the full experience of living through the pandemic. However, since this work was conducted in parallel with studies in older adults, people with mental health conditions, parents of young children and many others, the more general overlapping themes will be presented in future papers.

Our findings support large-scale quantitative surveys, but build on these by providing valuable depth and context, as well as factors that could potentially mitigate some of the negative impacts. The strength of qualitative research is that is can provide rich insights into people’s lived experience not attainable through quantitative methods. Interviews were conducted via video call or telephone, so could have missed some of the non-verbal cues and ability to build rapport and trust between participant and interviewer in person (Knox & Burkard, 2009). However, pandemic restrictions meant that in-person interviewing was not feasible, participants could choose telephone or video and the length and content of interviews suggest that participants still felt able to have an authentic discussion. In addition, it is also feasible that some people would be willing to share more over the phone. The interviews were conducted during a period when shielding was advised, and views might change as restrictions and guidelines change. However, we did capture anxieties about the future and consideration of how participants would feel when/if rules were relaxed. In addition, work conducted on previous pandemics helps shape the understanding of, and response to, future pandemics, so building this evidence base is important.

## Conclusion

The findings of this study suggest that living with a LTC during the COVID-19 pandemic had a significant impact on mental health and well-being, but also on attitudes towards physical health and use of health services. Our study highlights the interconnectedness of physical and mental health and illustrates the importance of managing stressors, and supporting health-preserving behaviours like symptom presentation and help seeking. Further, as people with LTCs are the most likely to have to experience longer periods of isolation during pandemics, whether enforced or self-directed as a protective strategy, there is a need for greater focus on how to ensure they are adequately supported both practically and socially.

## Data Availability

Data are qualitative interview transcripts and are not publically/freely available

## Funding statement and acknowledgements

The COVID-19 Social Study was funded by the Nuffield Foundation [WEL/FR-000022583], but the views expressed are those of the authors and not necessarily the Foundation. The study was also supported by the MARCH Mental Health Network funded by the Cross-Disciplinary Mental Health Network Plus initiative supported by UK Research and Innovation [ES/S002588/1], and by the Wellcome Trust [221400/Z/20/Z]. DF was funded by the Wellcome Trust [205407/Z/16/Z]. The funders had no final role in the study design; in the collection, analysis and interpretation of data; in the writing of the report; or in the decision to submit the paper for publication. All researchers listed as authors are independent from the funders and all final decisions about the research were taken by the investigators and were unrestricted.

The researchers are grateful for the support of a number of organisations with their recruitment efforts including: the UKRI Mental Health Networks, Alzheimers’s Society, University of Hertfordshire, NCRI Consumer forum, Third Aid Project, Yorkshire Cancer Community, Asian Women Cancer Group, Asthma UK. We would like to thank the COVID-19 Social Study team for ongoing discussion about analyses and Dr Rana Conway and Sara Esser for conducting some of the interviews.

**Funding acquisition:** Daisy Fancourt **Conceptualization:** Daisy Fancourt, Alex Burton, **Formal analysis:** Anna Roberts, Alison McKinlay **Writing - Original Draft** – Abi Fisher, Alison McKinlay, Alex Burton, Anna Roberts **Writing review and editing**: Abi Fisher, Alison McKinlay, Alex Burton, Daisy Fancourt, Anna Roberts

## Supplementary Material: Interview Topic Guide

*[Each section contained multiple prompts removed here to reduce word limit/length]*

### Ask to describe ‘normal life’

#### UNDERSTANDING AND ADHERENCE TO GUIDELINES

- **At the moment, are you self-isolating (how long for, reasons for this) a key worker, working but not a key worker, social distancing/ ‘staying at home’**
- **What do you understand by the ‘social distancing’ advice that is being given – what does it mean to you?**
- **Have you been able to stick to the social distancing advice that has been given to your group? Please tell us about why/ why not?**

#### LONG TERM CONDITION/CANCER

- **How has Covid-19 had an impact on the [long term condition/cancer]**

#### SOCIAL LIFE

- **How would you describe your social life before the Covid-19 pandemic?**
- **How would you describe your social life now that social distancing measures have been brought in because of Covid-19? Please tell us about this**

#### MENTAL HEALTH

- **How do you feel about the changes that have been brought about by Covid-19?**
- **Have they had any impact on your mental health or wellbeing? Please tell us about these**
- **Have you been doing/ planning anything to help with this?**
- **Why are you doing/ not doing these things?**

#### PROSPECTION

- **Has the pandemic meant that you have any worries for the future?**
- **How are these different from the worries you had before?**
- **Will this change the way you live your life in future?**
- **Has this changed any of your priorities for the future?**

